# Economic determinants of county-level mental health – United States, 2019

**DOI:** 10.1101/2024.03.08.24303977

**Authors:** Michele L.F. Bolduc, Parya Saberi, Torsten B. Neilands, Carla I. Mercado, Shanice Battle Johnson, Zoe. R.F. Freggens, Desmond Banks, Rashid Njai, Kai McKeever Bullard

## Abstract

A better understanding of whether and how economic factors impact mental health can inform policy and program decisions to improve mental health. This study looked at the association between county-level economic factors and the prevalence of self-reported poor mental health among adults in US counties in 2019, overall and separately for urban and rural counties. General dominance analyses were completed to rank-order the relative importance of the selected variables in explaining county prevalence of adults reporting > 14 poor mental health days in the last 30 days (“poor mental health”). The highest weighted variables were assessed for the statistical significance of their relationships with county-level poor mental health through multiple linear regression. Across all models, the four highest-ranked economic factors were household income, receipt of Supplemental Security Income (SSI), population with a college degree, and receipt of Supplemental Nutrition Assistance Program (SNAP) benefits. The overall, rural, and urban models explained over 69% of the variation in poor mental health prevalence between counties. Urban and rural models also showed notable differences in the relationship between poor mental health and median home value and population with public insurance. The findings from this study indicate a significant association between several economic factors and poor mental health, which may inform decision makers in addressing mental health in the US.

## Introduction

Mental health is a key component of overall health and well-being, and mental health conditions are common in the United States.^1^ The prevalence of any mental, behavioral, or emotional disorder among US adults has been increasing since 2008, reaching 20.6% by 2019.^2^ An estimated 9.2% of adults have received mental health treatment in the last 12 months^3^, but almost half (43.8%) of adults who needed treatment did not receive it.^2^ Poor mental health, which includes stress, depression, and problems with emotions – is a risk factor for a range of chronic diseases and injuries, from stroke to diabetes.^4–6^ Poor mental health is also costly for the economy.^7^ Major Depressive Disorder alone cost the US an estimated $326 billion in 2018, a 37.9% increase over the costs in 2010, primarily due to increasing workplace costs.^8^ However, many factors that affect mental health could be prevented or mitigated to improve overall public health.

The biopsychosocial model of mental health acknowledges the role that social, economic, and political factors play in the development and worsening of poor mental health.^9^ In the free market-oriented American economy, economic factors, including employment status and income, play a key role in our lives. One survey found that among people in the US, the top personal stressors were work (64% of respondents) and money (60%); 46% indicated the economy was a significant source of stress.^1^ Exposure to adverse conditions, such as material hardship, “undoubtedly leads to stress and known psychological and physiological stress responses” that increase the risk of mental health conditions.^9^ These issues may be exacerbated on a population level in difficult economic times, with higher rates of suicide, homicide, substance abuse, and psychiatric disorders noted during financial crises.^10–13^ An exploration of the economic factors that impact mental health could inform possible public health interventions.

Improving population mental health and reducing adverse mental health outcomes requires a shifting of focus from individual treatment – which is costly to patients and the healthcare system – to population-level or systems-based approaches. Specifically, there is a need to move upstream from the more proximal social determinants of health to the structural drivers of those determinants.^14–16^ Structural drivers relate to how policy and economics shape the distribution of resources; changes at the structural level have the potential for greater impacts on population health and health equity.^14,17^ Upstream interventions in policy, programs, and practice place the responsibility of health on decision-makers to undertake broader systemic change rather than focus on individual choice and behavior^9^, which could have impacts across the life course and across generations.^18^

Data demonstrating whether economic factors may be associated with mental health can inform policy and program decisions. This study, therefore, examines the association between county-level economic factors and the prevalence of self-reported poor mental health among adults in US counties in 2019.

## Methods

This analysis is a cross-sectional study of the association between county-level economic determinants of health (SSDOH) variables (e.g., county Gross Domestic Product [GDP], income inequality, housing affordability) and self-reported poor mental health prevalence for US counties in 2019. Data for 2019 were used to get a baseline understanding of the economic factors associated with mental health prior to the start of the COVID-19 pandemic.

The dependent variable of self-reported poor mental health comes from Centers for Disease Control and Prevention (CDC) PLACES data (www.cdc.gov/places), which produces small area estimates using data from the CDC’s Behavioral Risk Factor Surveillance System (BRFSS). The BRFSS (www.cdc.gov/brfss) is a telephone survey through which states collect information on health-related risk behaviors and health conditions, with over 400,000 adult participants each year. The survey asks the question, “Now thinking about your mental health, which includes stress, depression, and problems with emotions, for how many days during the past 30 days was your mental health not good?”^19^ BRFSS defines poor mental health as responses indicating >14 poor mental health days over the last 30 days. PLACES uses the BRFSS data to generate county-level prevalence estimates of poor mental health for all US states and the District of Columbia, using a multilevel statistical model that considers individual-level demographic data from BRFSS, county-level percentage of adults below 150% of the federal poverty level from the American Community Survey (ACS), and state- and county-level random effects.^20^ Estimates for poor mental health prevalence are available for 3,121 counties for 2019, with 23 missing counties in New Jersey (not available from BRFSS) and Alaska (missing the newer Chugach and Copper River Census Areas).

Economic SSDOH variables originate from the US Bureau of Economic Analysis (BEA) and from American Community Survey (ACS) five-year estimates for 2019 (S1 Table). Variables were chosen to measure economic factors (relating to the production, distribution, and consumption of goods and services) meaningful at the county level. Business variables from BEA were selected to measure current and changing county-level economic activity, including real GDP, 10-year change in GDP, and 10-year change in county GDP. Business variables from BEA are not available for 51 jurisdictions in Virginia due to combined independent city/county estimates, nor for two counties in Hawaii (Maui and Kalawao) due to combined county estimates. Employment variables covered a range of work-related issues, including unemployment rate, employed but under the Federal Poverty Level (known throughout this paper as ‘working poverty rate’), mean usual hours worked over the last 12 months, employees working from home, and mean travel time to work. Income and wealth variables include median individual earnings, median household income, population with a college degree, two measures of inequity (Gini Index and the gender pay gap), percent of the population with public assistance income (Social Security, Supplemental Security Income [SSI], and cash public assistance), as well as homeownership rate as a significant component of wealth building. Several variables were included to cover key expenses – two housing affordability variables (for homeowners, population with Selected Monthly Owner Costs as a Percentage of Household Income [SMOCAPI] costs of at least 30%, and, for renters, population with Gross Rent as a Percent of Household Income [GRAPI] costs of at least 30%), a food security variable (percent receiving Supplemental Nutrition Assistance Program or SNAP benefits), and two health insurance coverage variables (percent uninsured and percent with public health insurance coverage). Two more variables were added to capture county-level economics, including median home value and 10-year population change.

Our first step was to use PLACES data to map the prevalence of poor mental health at the county level for 2019 in RStudio version 4.2.1 (R Foundation for Statistical Computing, Vienna, Austria). Stata version 17.0 (StataCorp LP, College Station, TX, USA) was used to complete the following analyses for all counties and for counties classified as either urban or rural based on the 2013 USDA Rural-Urban Continuum Codes. The median and Interquartile Range (IQR) for each variable were described across all counties in the datasets. Next, variable clustering was used to remove two highly collinear (>0.5 Spearman coefficient) and redundant (based on our understanding of the literature) variables from our analysis: median earnings (keeping median household income) and change in real GDP (keeping change in GDP). General dominance analyses were then completed using the Stata package *domin* with all the remaining economic variables to rank-order the relative importance of the variables in explaining county-level poor mental health.^21^ This method finds the proportion of the total R^2^ accounted for by each variable based on the average R^2^ across all possible subsets of regression models. Using a scree plot (S1 Fig), the most important (highest weighted) variables were identified from the dominance analyses based on dominance weights above the “elbow” of the graph, retaining eleven important variables. The eleven variables were then assessed for the statistical significance of their relationships with county-level poor mental health through multiple linear regression.

## Results

Fig 1 shows the prevalence of poor mental health across all counties in 2019, with a mean of 16.0% and a median of 15.8% (IQR: 14.1 – 17.8%). County prevalence ranged from 9.7% (Falls Church, VA) to 26.3% (East Carroll Parish, LA). Across the US, prevalence was higher in Appalachia and the Deep South and parts of Alaska, Montana, South Dakota, and the Southwest. Lower prevalence was found in the Upper Midwest.

**Fig 1.**
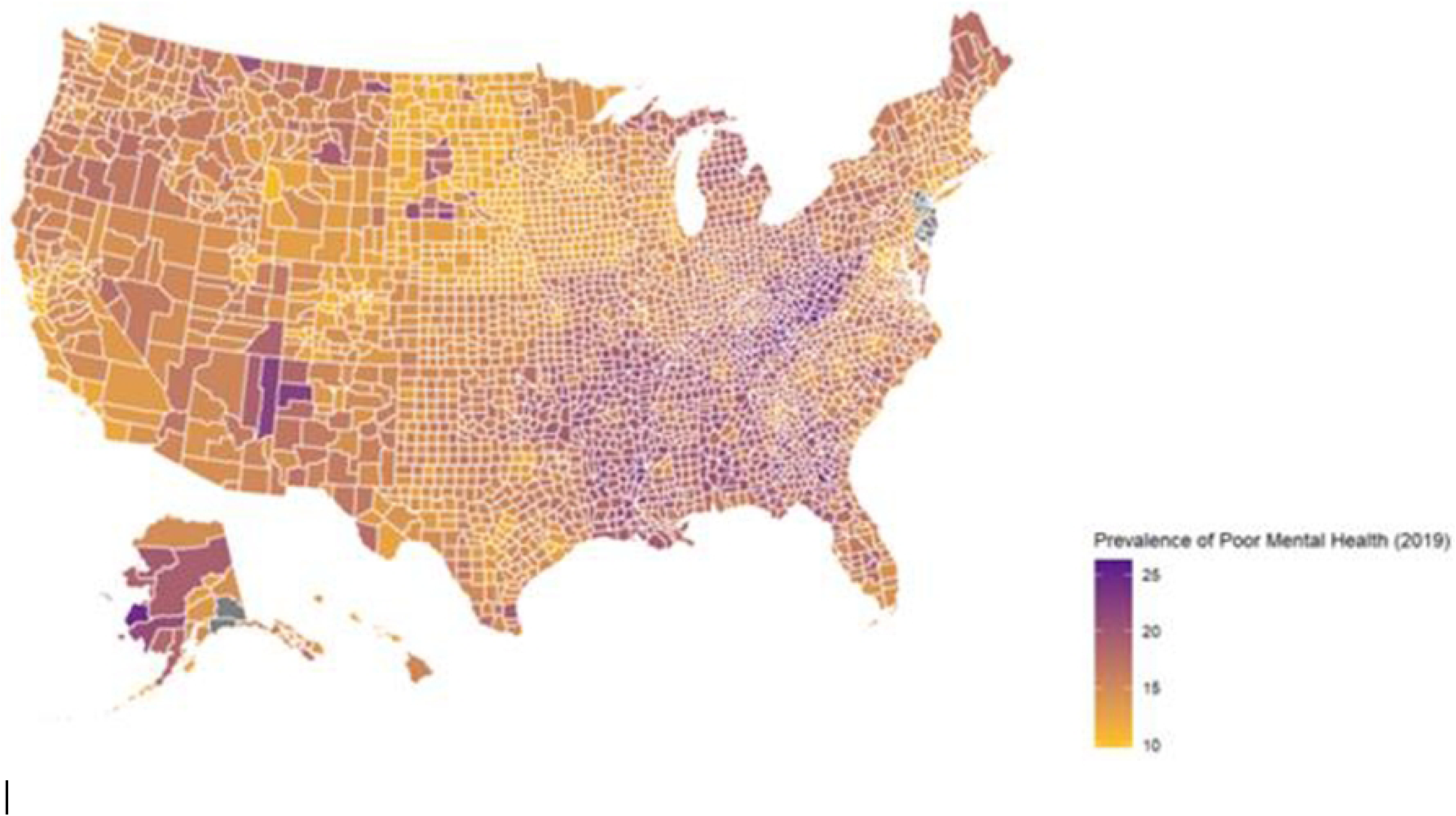
Estimated Prevalence of Poor Mental Health by County, United States, 2019. Data come from CDC PLACES, which is modeled from the CDC Behavioral Risk Factor Surveillance System (BRFSS). BRFSS asks the question, “Now thinking about your mental health, which includes stress, depression, and problems with emotions, for how many days during the past 30 days was your mental health not good?” Self-reported poor mental health is defined as responses of >14 poor mental health days over the last 30 days.

Median and IQR values for each economic variable in 2019 are listed in Table 1. Table 2 shows the most important economic variables for county-level poor mental health identified using the dominance analysis based on standardized dominance weights. The top variables were median household income, percent of households with SSI, percent of the population 25 years or older with a college degree, percent of households receiving SNAP benefits in the last 12 months, and percent of the population with public health insurance. The top results were similar for urban counties, although unemployment had lower standardized dominance weights. For rural counties, the results were similar, but travel time from work ranked higher.

**Table 1.** Summary Statistics for Economic Determinants of Health Variables Used to Analyze County-Level Association Between Economic Factors and Estimated Prevalence of Poor Mental Health, United States, Overall and by Urban/Rural Classification, 2019.

| Factor | Variable | 2019<br>Median<br>[IQR] |  |  |
| --- | --- | --- | --- | --- |
|  |  | Overall<br>N=3,121 | Urban<br>n=1,145 | Rural<br>n=1,976 |
| Business | 10-year percent change in Gross Domestic Product (GDP), 2010 – 2019 (%) | 28.2%<br>[15.3 – 42.8%] | 35.6%<br>[22.6 – 49.2%] | 24.6%<br>[10.9 – 39.1%] |
| | Real GDP, in thousands of chained dollars (\$) | \$946,103<br>[\$361,013 – \$2,691,786] | \$4,018,925<br>[\$1,023,404 – \$10,900,000] | \$595,270<br>[\$263,664 – \$1,295,594] |
|  | Change in Real GDP, 2010 – 2019 (%) | 9.7%<br>[-1.8 – 22.3%] | 14.1%<br>[3.6 – 26.3%] | 6.4%<br>[-4.38 – 19.5%] |
| Employment | Unemployment rate, ≥ 16 years old (%) | 4.9%<br>[3.6 – 6.5%] | 4.9%<br>[3.9 – 6.1%] | 5.0%<br>[3.4 – 6.8%] |
|  | Employed but under the Federal Poverty Limit (FPL) (%) | 6.8%<br>[5.1 – 8.9%] | 6.0%<br>[4.4 – 7.9%] | 7.3%<br>[5.5 – 9.3%] |
|  | Mean usual hours per week worked in the past 12 months for workers 16-64 years old | 39.3 hours<br>[38.5 – 40.2 hours] | 39.0 hours<br>[38.3 – 39.7 hours] | 39.6 hours<br>[38.6 – 40.7 hours] |
|  | Employees ≥ 16 years old working from home (%) | 4.5%<br>[3.2 – 6.3%] | 4.5%<br>[3.4 – 5.8%] | 4.4%<br>[3.0 – 6.7%] |
|  | Mean travel time to work (minutes) | 23.7 minutes<br>[19.8 – 27.5 minutes] | 25.7 minutes<br>[22.4 – 29.5 minutes] | 22.4 minutes<br>[18.4 – 26.0 minutes] |
| Income and Wealth | Median earnings (\$) | \$32,607<br>[\$30,660 – \$36,916] | \$35,963<br>[\$32,175 – \$40,070] | \$31,738<br>[\$29,801 – \$34,995] |
| | Median household income (\$10,000 increments) | \$5.2<br>[\$4.4 – \$6.0] | \$5.8<br>[\$5.1 – \$6.8] | \$4.9<br>[\$4.2 – \$5.5] |
|  | Population ≥ 25 years old with a college degree (%) | 19.5%<br>[15.3 – 25.8%] | 24.8%<br>[18.6 – 33.1%] | 17.8%<br>[14.3 – 22.1%] |
|  | Gini Index of income inequality (index 0-1) | 0.44<br>[0.42 – 0.47] | 0.44<br>[0.42 – 0.47] | 0.44<br>[0.42 – 0.47] |
|  | Female pay as a percentage of male pay (gender pay gap) (%) | 76.5%<br>[71.4 – 81.8%] | 78.0%<br>[73.4 – 82.3%] | 75.8%<br>[70.2 – 81.3%] |
|  | Households with Social Security income (%) | 37.4%<br>[32.8 – 41.8%] | 33.8%<br>[29.4 – 37.9%] | 39.2%<br>[35.3 – 43.3%] |
|  | Households with Supplemental Security Income (SSI) (%) | 5.5%<br>[4.1 – 7.3%] | 5.1%<br>[4.0 – 6.6%] | 5.8%<br>[4.2 – 8.0%] |
|  | Households with public cash assistance (%) | 1.9%<br>[1.3 – 2.8%] | 1.9%<br>[1.4 – 2.6%] | 1.9%<br>[1.3 – 2.9%] |
|  | Homeownership (%) | 72.9%<br>[67.6 – 77.2%] | 71.7%<br>[64.6 – 77.0%] | 73.4%<br>[68.9 – 77.2%] |
| Expenses | Prevalence of units with a mortgage with Selected Monthly Owner Costs as a Percentage of Household Income (SMOCAPI) of 30% or more (%) | 24.6%<br>[20.9 – 29.4%] | 24.2%<br>[21.2 – 28.5%] | 24.8%<br>[20.6 – 29.8%] |
|  | Prevalence of rent-paying units with Gross Rent as a Percentage of Household Income (GRAPI) of 30% or more (%) | 44.3%<br>[38.1 – 49.5%] | 46.6%<br>[42.3 – 50.8%] | 42.4%<br>[35.8 – 48.4%] |
|  | Households with Supplemental Nutrition Assistance Program (SNAP) benefits in the past 12 months (%) | 11.9%<br>[8.3 – 16.2%] | 11.1%<br>[8.0 – 14.6%] | 12.6%<br>[8.5 – 17.1%] |
|  | Population 19-64 years old without health coverage (%) | 8.7%<br>[5.8 – 12.1%] | 8.0%<br>[5.3 – 10.8%] | 9.1%<br>[6.1 – 13.2%] |
|  | Population with public health insurance coverage alone (Medicaid, Medicare, Veterans Administration [VA]) (%) | 39.4%<br>[33.2 – 45.3%] | 35.6%<br>[30.3 – 41.3%] | 41.7%<br>[35.7 – 47.3%] |
| Neighborhood | Median home value, in \$10,000 increments (\$) | \$12.7<br>[\$9.8 – \$17.4] | \$16.7<br>[\$13.0 – \$22.6] | \$11.0<br>[\$8.8 – \$14.4] |
|  | 10-year population change (%) | -0.07%<br>[-3.6 – 5.2%] | 4.3%<br>[-0.35 – 10.5%] | -1.8%<br>[-4.9 – 2.0%] |
| Dependent Variable | Prevalence of > 14 poor mental health days of the last 30 days, age-adjusted (%) | 15.8%<br>[14.1 – 17.8%] | 15.3%<br>[13.7 – 16.9%] | 16.1%<br>[14.4 – 18.4%] |

NB: The three Business variables are missing 2 counties in Hawaii and 51 counties in Virginia (Overall Analytic N=3,068; Urban n=1,109; Rural n=1,959).
\* ACS = American Community Survey, BRFSS = Behavioral Risk Factor Surveillance System

**Table 2.** Most-to-Least Important County-Level Economic Variables Contributing to County Prevalence of Poor Mental Health Based on Dominance Analysis Ranked by Standardized Dominance Weights, Overall and by Urban/Rural Classification, United States, 2019.

| 2019 Dominance Analysis Rankings and Weights |  |  |  |  |  |
| --- | --- | --- | --- | --- | --- |
| Overall<br>N=3,068 | Weights | Urban<br>n=1,109 | Weights | Rural<br>n=1,959 | Weights |
| 1. Household income | 0.1155 | 1. Household income | 0.1203 | 1. Household income | 0.1180 |
| 2. Receipt of SSI | 0.1111 | 2. Receipt of SSI | 0.1131 | 2. College degree | 0.1091 |
| 3. College degree | 0.0989 | 3. College degree | 0.0918 | 3. Receipt of SSI | 0.1032 |
| 4. Receipt of SNAP | 0.0943 | 4. Receipt of SNAP | 0.0827 | 4. Receipt of SNAP | 0.0897 |
| 5. Public insurance | 0.0834 | 5. Public insurance | 0.0789 | 5. Work travel time | 0.0822 |
| 6. Unemployment | 0.0701 | 6. Social Security | 0.0757 | 6. Public insurance | 0.0789 |
| 7. Work from home | 0.0670 | 7. Home values | 0.0746 | 7. Unemployment | 0.0762 |
| 8. Work travel time | 0.0579 | 8. Work from home | 0.0488 | 8. Work from home | 0.0744 |
| 9. Home values | 0.0576 | 9. Working poverty | 0.0485 | 9. Working poverty | 0.0496 |
| 10. Social Security | 0.0549 | 10. Unemployment | 0.0431 | 10. Home values | 0.0388 |
| 11. Working poverty | 0.0527 | 11. Work travel time | 0.0315 | 11. Social Security | 0.0385 |
| 12. Gini Index | 0.0283 | 12. Population change | 0.0298 | 12. Gini Index | 0.0345 |
| 13. Uninsured | 0.0192 | 13. SMOCAP≥30% | 0.0235 | 13. GRAP≥30% | 0.0230 |
| 14. Population Change | 0.0160 | 14. Real GDP | 0.0233 | 14. Hours worked | 0.0198 |
| 15. Real GDP | 0.0139 | 15. Uninsured | 0.0213 | 15. Uninsured | 0.0157 |
| 16. GRAP≥30% | 0.0123 | 16. Homeownership | 0.0186 | 16. Cash assistance | 0.0108 |
| 17. Homeownership | 0.0111 | 17. Gini Index | 0.0185 | 17. Homeownership | 0.0104 |
| 18. Hours worked | 0.0102 | 18. Change GDP | 0.0172 | 18. SMOCAP≥30% | 0.0102 |
| 19. Cash assistance | 0.0090 | 19. Gender pay gap | 0.0171 | 19. Real GDP | 0.0065 |
| 20. SMOCAP≥30% | 0.0086 | 20. Cash assistance | 0.0089 | 20. Population change | 0.0052 |
| 21. Change GDP | 0.0054 | 21. Hours worked | 0.0070 | 21. Change GDP | 0.0029 |
| 22. Gender pay gap | 0.0027 | 22. GRAP≥30% | 0.0056 | 22. Gender pay gap | 0.0025 |

\* SSI = Supplemental Security Income, SNAP = Supplemental Nutrition Assistance Program, GDP = Gross Domestic Product, GRAP = Gross Rent as a Percent of Household Income, SMOCAP = Selected Monthly Owner Costs as a Percentage of Household Income

In the overall model using our top eleven variables identified in the scree plot, county-level measures significantly positively associated with the prevalence of poor mental health included: unemployment, working poverty rate, mean travel time to work, households with Social Security, households with SSI, and households with SNAP benefits (Table 3). County-level measures significantly inversely associated with poor mental health prevalence were employees working from home, median household income, and percent of the population with a college degree. The largest coefficients were in median household income, percent of the population working from home, and population receiving SSI. The variables in the overall model explained 70% (Adjusted R^2^ = 0.7001) of the variation in poor mental health prevalence between counties.

**Table 3.**
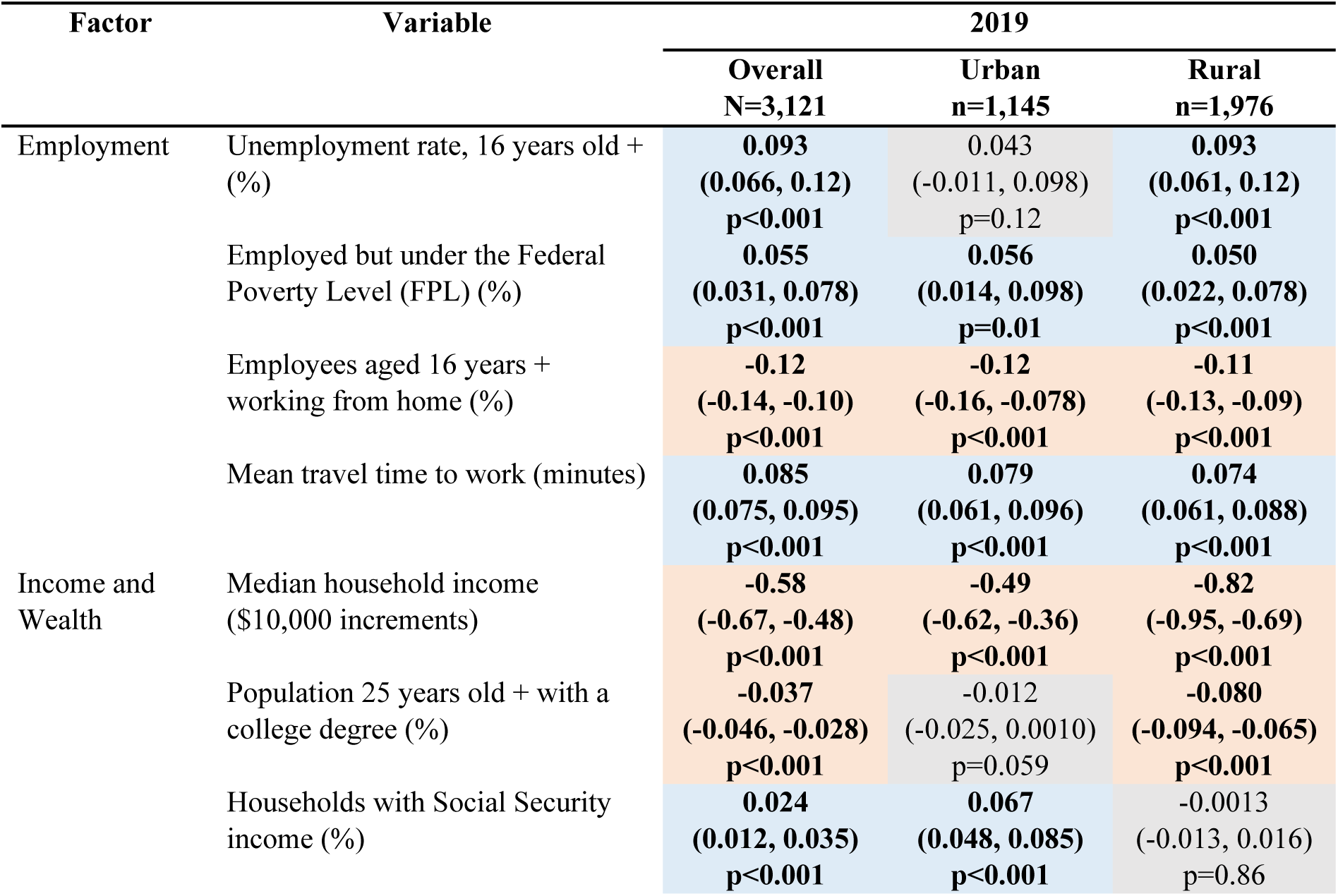
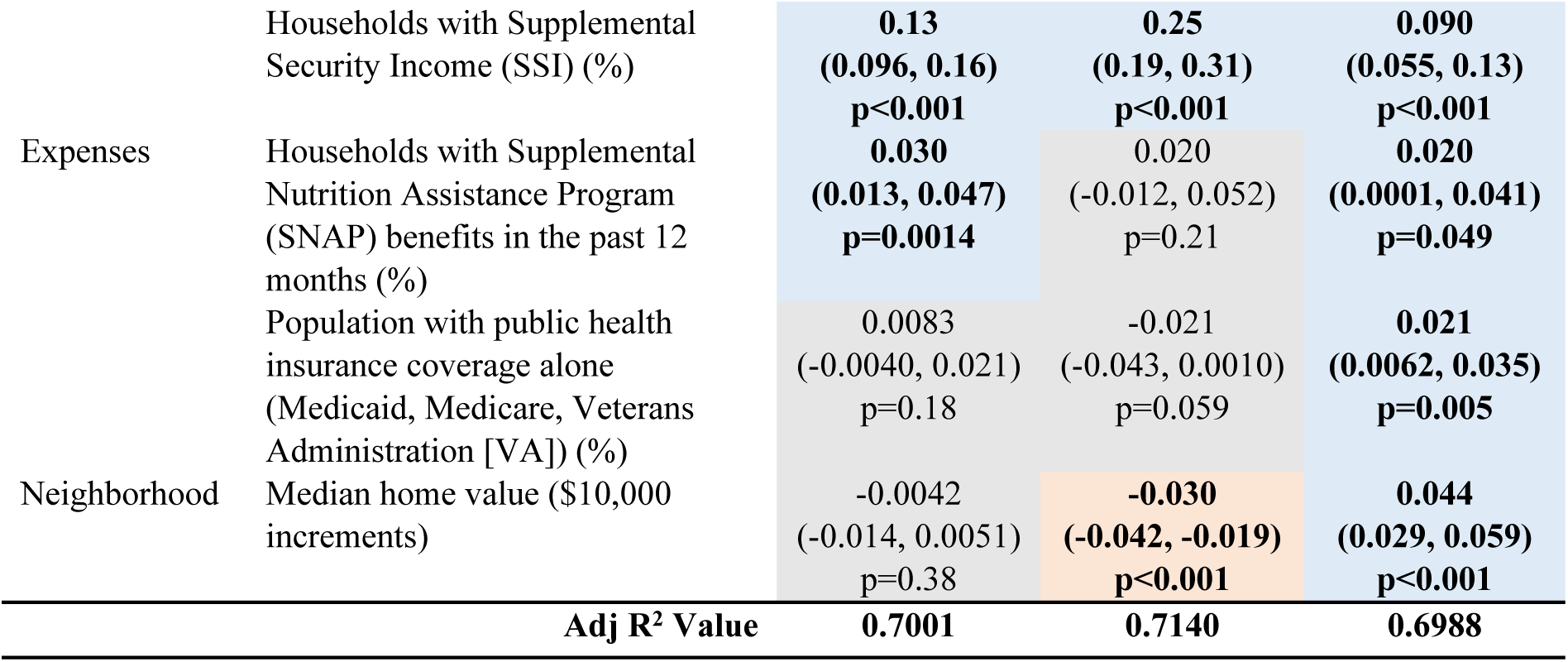
Beta Coefficients, 95% Confidence Interval, and Statistical Significance for County-Level Economic Variables Using Linear Regression with Prevalence of Poor Mental Health as the Dependent Variable, Overall and by Urban/Rural Classification, United States, 2019. Blue-filled cells indicate a positive association between the variable and the dependent variable; red-filled cells indicate a negative association; greyed out cells indicate the variable was not significant.

For urban counties, the unemployment rate, population with a college degree, and households with SNAP benefits were not statistically significantly associated with poor mental health prevalence. However, median home values were significantly inversely associated with the prevalence of poor mental health. In the rural model, nine county-level economic measures used were statistically significantly associated with poor mental health prevalence; households with Social Security and households with SNAP benefits were not significant. In contrast to urban counties, median home values in rural counties were significantly positively associated with poor mental health. The adjusted R^2^ values indicate that the urban and rural models explained 71.4% and 69.9% of the variation in poor mental health, respectively.

## Discussion

We identified the top eleven important economic determinants of self-reported poor mental health at the US county level; these variables explained over 70% of the variation in poor mental health between counties in 2019. These eleven explanatory variables associated with poor mental health were consistent in urban and rural models with some variation in the ranking. Across all models, the four highest-ranked economic factors were related to income and expenses: household income, receipt of SSI, population with a college degree, and receipt of SNAP benefits.

Based on our findings, median household income was the most important factor associated with county-level poor mental health in both urban and rural areas. Our results show that higher median income in a county is associated with a lower prevalence of poor mental health. Income is a key determinant of health because it provides households with the resources necessary for health and wellbeing, including basic needs like nutritious food, safe housing, and health care services and other resources like higher education.^22^ Low income puts children and adults at higher risk for stress, worse physical health, exposure to adverse conditions in early life, and exposure to violence and crime, any of which could trigger mood and anxiety disorders.^23–25^ A reduction in or loss of income may be particularly harmful to mental health.^23,26–27^ In addition, there is ample evidence that interventions aimed at reducing poverty, such as through cash support, have a positive effect on mental health, particularly when the support lifts someone out of poverty.^23,26^

One type of income support, SSI benefits, was also significant in the model. The results showed that as the percentage of the population with SSI benefits increased, so too did the prevalence of poor mental health. SSI provides fixed, monthly supplemental financial assistance to people who are: under 65 years old who are blind or have a disability (including mental disorders) that makes it difficult to perform substantial gainful activity or over 65 years old without a disability who live on a low income. Among the 72% of SSI recipients who are under 65 years old, six out of 10 were receiving SSI due to a mental health disorder.^28^ In addition, adults with disabilities or who have low incomes are more likely to report frequent mental distress than those without disabilities or with higher incomes.^29^ The association between receipt of SSI and prevalence of poor mental health may be due to the higher rate of mental health disorders among SSI recipients than among the general public; however, it may also suggest that the program on its own may not be able to mediate the impact that structural and social inequities have on the number of poor mental health days experienced by people with disabilities and/or low incomes.

Similarly, a higher number of SNAP recipients in a county was associated with higher prevalence of poor mental health, with a stronger relationship in the rural than in the urban model. To be eligible for SNAP benefits, households must meet certain gross and net income and resource limits, making SNAP a proxy for low-income households. Under special SNAP rules, recipients with disability income (e.g., SSI) only need to meet net income limits. The positive relationship between SNAP benefits and poor mental health may reflect higher prevalence of mental health conditions among recipients. An estimated 28% of non-elderly SNAP recipients have a disability (which could include mental health conditions); of those with a disability, almost 60% receive social security benefits.^30^ Though SNAP benefits may have a positive impact on public health^31^, the results of this model suggest that SNAP benefits may not be able to make up for the financial gaps people with low incomes and/or disabilities may have and the impacts of these financial shortfalls on mental health.

Although the percentage of the population with public health insurance coverage was a top factor in the dominance analysis for all models, it was only statistically significant (positive) in the rural counties analysis. This suggests that income-based health insurance coverage may play a bigger role in poor mental health prevalence in rural areas than urban ones. Like SSI and SNAP households discussed previously, the positive relationship between public health insurance coverage and poor mental health may indicate that access to public insurance may not mediate the impact of having a low income, and this appears to be particularly true for rural areas.

The significance of having a college degree for lower prevalence of poor mental health may be related to higher wages associated with greater educational attainment^32^ and the increased potential to build wealth. College education also increases opportunities for jobs with benefits like health coverage and increased access to health-promoting knowledge and resources, resulting in lower rates of death and longer lives.^9^ Lower levels of educational attainment are associated with increased risk of some psychiatric diagnoses like Major Depressive Disorder and Generalized Anxiety Disorder^33^ and higher rates of suicide.^34^ Conversely, poor mental health is associated with higher risk of early termination of education, limiting educational attainment.^35^

In addition to the income and expense variables, we found that as mean travel time to work increased, the prevalence of poor mental health increased, which could be explained as a function of the increased time spent making driving-related decisions, leading to higher stress levels.^36^ The results showed that the percentage of employees working from home was inversely associated with poor mental health, which also may be related to commute time and other aspects of remote work that support mental well-being, such as fewer distractions, a more comfortable environment, and more time to spend with family or cook meals.^37^

An unexpected result was the null finding for median home values across all US counties, yet significant associations for both the urban and rural models – but in opposite directions. Median home value in urban counties had a significant inverse relationship with poor mental health. This is consistent with other studies like those on the Moving to Opportunity for Fair Housing Demonstration Program that have shown that higher income and higher opportunity neighborhoods may improve mental and physical health, especially for adults^38–40^, possibly relating to less exposure to neighborhood level stressors like crime^40^ and greater access to health-promoting resources like high-quality medical providers^41^ and recreation facilities and amenities in good condition^42^ than are available in poorer neighborhoods. However, higher home values in rural areas are associated with higher prevalence of poor mental health – though the importance of median home values ranked lower in rural areas than in urban areas. Lower wages, fewer opportunities for high-paying jobs, and slower recovery of employment since the Great Recession^43^ in rural places may indicate greater financial insecurity among rural counties, including among homeowners. The contrasting relationships to poor mental health in urban and rural counties support the need to examine associations with urban and rural economic factors separately.

While individual- and clinical-level interventions to address these economic drivers exist, such as connecting patients receiving mental health care with economic resources to ease financial burdens, these types of interventions are not sufficient.^9^ Population health improves as income increases, particularly for populations in the lowest income brackets.^23,26^ Interventions that systematically address poverty and the financial burden of basic needs like health insurance and food may have a greater impact on health in the US than a narrow focus on individual interventions.^17^ Addressing the upstream economic drivers of poor mental health may also have cascading impacts on a range of other chronic health outcomes that share the same drivers, from mortality to substance abuse.^44–47^

Structural interventions relating to household income that may have a positive effect on county-level mental health include expanding programs for federal income subsidies^48^; ensuring income support for households during difficult financial times, such as unemployment^49–50^; establishment of a universal basic income^51^; increasing federal, state, or local minimum wages^52–53^; offering additional tax breaks or credits for low-income households^54–55^; and enacting policies which promote equitable access to higher-paying employment opportunities.^56^ Programs like SSI that provide financial assistance to people with poor mental health who may have challenges maintaining stable work and health insurance for treatment could positively impact mental health. Similarly, policies and programs that support people with low incomes so they can afford necessary expenses like housing and food may have a beneficial impact on mental health, beyond the original intentions of the program.^57–58^ Additional interventions that might benefit population mental health include increasing educational opportunities, such as high school completion initiatives, low-cost technical college programs, and college tuition assistance^59–60^; expanding access to healthcare through subsidized or universal health insurance coverage^61–63^; and thoughtful implementation of policies supporting work from home.^64^

Future analyses could evaluate which approaches are most likely to improve mental health. However, economic considerations may be necessary but insufficient to comprehensively address the persistence of poor mental health in the US. A systems approach would help understand the most effective ways to improve mental health. Impactful changes in one place can have cascading effects and create systemic change.^18^ Assessment of the mental health consequences of any new policy or policy change could be standard practice, in line with a Health in All Policies approach.^65^ However, the full effects of large-scale, structural, and systemic changes may not be seen for many years.^7^

Additional research could clarify the relationships between economic factors and mental health at the individual level; however, more SSDOH data (such as standardized data in electronic health systems^66^) are needed to capture how mental health may be influenced by SSDOH on an individual basis. Relatedly, the importance of each economic variable would be expected to vary depending on membership in various intersecting minority groups (e.g., age, sex, sexual orientation and gender identity, race, ethnicity, and nativity). Past and present discrimination against members of minority groups has had lasting economic consequences for group members^67^, and these inequities are not fully reflected in existing datasets. Future research could continue to explore the relationships between economic factors and health inequities, as well as the most effective change points for interventions to improve the association between economic factors and a range of health outcomes.

Our study had three main limitations. First, we conducted an ecological study to assess county-level associations, limiting our ability to infer causality. Second, this study carries forward the limitations of our original data sources. For example, the mental health prevalence estimates use data from BRFSS, which uses self-reported answers to a single question to assess mental health status. Third, reduced sample sizes in the urban- and rural-specific models may have impacted our findings.

Nevertheless, this study provides a unique list of county-level economic factors that may influence mental health status that could be used as a starting point for further analysis. It provides a baseline analysis to understand economic factors related to mental health prior to the major economic disruptions associated with the COVID-19 pandemic, which worsened mental health challenges.^68–70^ Finally, it applies dominance analysis techniques to go beyond the identification of potential drivers to rank these drivers in order of importance, which can inform the prioritization of policies and programs to improve mental health status.

## Conclusions

This study provides important data to researchers and policymakers on the association of economic factors and mental health. This study found that higher county-level access to resources, including income, education, and health insurance, is associated with a lower prevalence of poor mental health. In our market-based economy, economic factors shape the distribution of resources and opportunities needed to maintain mental well-being and these distributions are geographically uneven across the US. Additional research could explore additional economic variables as indicators and their relationship to specific mental health outcomes (e.g., depression); spatial trends in poor mental health over time; and possible mental health-specific population-level interventions and their effectiveness in improving mental health to improve public health and address health inequities.

## Data Availability

All data is available from public data sources, including the US Bureau of Economic Analysis (www.bea.gov/data), the US Census (data.census.gov), and CDC PLACES data (www.cdc.gov/PLACES).

## Acknowledgements

The findings and conclusions in this manuscript are those of the authors and do not necessarily represent the official position of the Centers for Disease Control and Prevention. The authors would like to recognize the CDC Health Equity Science Manuscript Fellowship in partnership with the University of California, San Francisco, for the dedicated training and collaboration that have contributed to this publication. This manuscript also benefitted from statistical consultation with the UCSF Clinical and Translational Science Institute.

## Supporting information

**S1 Fig. Scree Plot of Dominance Weights Identified by the Dominance Analysis of Economic Variables in Relation to Poor Mental Health.** The “elbow” on the plot at variable 12 (indicated in red) shows where the dominance weights drop off most steeply in value; values to the left of the elbow (11 total) were retained as significant for our multiple linear regression models.

**S1 Table. Economic Determinants of Health Variables Used to Analyze County-Level Association Between Economic Factors and Estimated Prevalence of Poor Mental Health and their Sources, United States, 2019.**

